# Prediction of New Coronavirus Infection Based on a Modified SEIR Model

**DOI:** 10.1101/2020.03.03.20030858

**Authors:** Zhou Tang, Xianbin Li, Houqiang Li

## Abstract

**BACKGROUND:** The outbreak of the new coronavirus infection in Wuhan City, Hubei Province in December 2019, poses a huge threat to China and even global public health security. Respiratory droplets and contact transmission are the main routes of transmission of new coronaviruses. Compared with SARS and Ebola viruses, new coronavirus infections are infectious during the incubation period. Traditional SEIR (susceptibility-exposure-infection-Removal) There are some differences in conditions for the prediction of the epidemic trend of new coronavirus infection. The outbreak of the new coronavirus infection coincided with the Spring Festival before and after the Chinese Spring Festival.It is necessary to make appropriate optimization and amendments to the traditional model to meet the actual evolution of the epidemic situation.

**METHODS:** The traditional SEIR model assumes that the virus-infected person is not infectious during the incubation period and that the infected person did not take isolation measures during the illness. The transmission of the new coronavirus no longer meets the basic assumptions of the classical kinetic system. Therefore, this article first establishes a modified SEIR model. Predict and analyze the changing trend of the epidemic situation, then estimate the parameters involved in the infection dynamics model, and then use Matlab to simulate the established dynamic equations based on public data and analyze the results. Recommendations for universal prevention and control of infectious diseases.

**RESULTS:** The first case of new coronavirus infection was confirmed in Wuhan on December 8, 2019. When Wuhan City took no action, assuming the average daily number of contacts per infected person k = 5, the number of infected persons will reach about 2,384,803 people; If wuhan adopts the measures of sealing the city on January 22, 2020, under the premise of k=2, the number of infected people decreases by 19,773 compared with that on January 23, and there is no significant change in the time when the number of infected people reaches the peak. Under the premise of k = 1, the number of infected persons was reduced by 14,330 compared with the closure on January 23, and the time to reach the peak of the number of infected persons was reduced by 2 days. If Wuhan City is closed for one day, the number of infected persons will increase from 106,145 to 130,626 under the premise of k = 2; the number of infected persons will increase from 74,369 to 92,010 under the premise of k = 1.

**CONCLUSIONS:** Comparing the number of confirmed diagnoses actually notified by the department with the number of infected people obtained from the simulation of the model, it can be seen that the city closure measures adopted by the Wuhan Municipal Government on January 23 and the first-level response measures adopted by the country are effective for the epidemic Prevention and control play a vital role. Wearing a mask when going out and avoiding close contact with people can effectively reduce the infection rate.

## INTRODUCTION

At the beginning of the outbreak of the new coronavirus infection, some cases were related to the South China Seafood Market in Wuhan, Hubei. At first, it was diagnosed as “unexplained pneumonia”, but it quickly spread to all parts of the country and parts of Southeast Asia, North America, and Europe. Wuhan City, Hubei Province, as a place of communication between the nine provinces, the large population movement, the influence of geographical factors, and the transmission characteristics of the new coronavirus have added many difficulties to the prevention and control of the epidemic. As of 13th February 2020, an outbreak of COVID-19 has resulted in 46,997 confirmed cases^1^. As of March 3, 2020, 5186 cases have been confirmed in South Korea, 1501 cases have been confirmed in Iran, 2036 cases have been confirmed in Italy, and 80,303 cases have been confirmed in China, of which 67,217 have been diagnosed in Hubei Province, accounting for 83.7%. In the early stage, understanding the dynamic mechanism of the spread of the new coronavirus is important to control the spread of the virus from person to person. About 5 million outflows occurred in Wuhan during the Spring Festival, about 70% of which went to other cities in Hubei Province. About 30% went to provinces, municipalities and municipalities across the country, which made it difficult for the infected people to trace back to the population. Except for Hubei Province, the newly confirmed cases in other parts of the country are generally stable, with a low case fatality rate of about 2% to 3%. The National Health and Health Commission has incorporated new coronavirus infection into Class B infectious diseases and adopted Class A prevention and control measures.

The source of the new coronavirus has not been determined so far. Various evidences show that the source of the infection of the new coronavirus comes from wild animals. It is most similar to a coronavirus isolated from bats. The intermediate host may be pangolin, but which kind of wild animal uncertain. The new coronavirus causes human-to-human transmission mainly by respiratory droplets, causing fever, cough, and shortness of breath. It is also infectious during the incubation period. In conclusion, considering transmissibility and fatality rate, 2019-nCoV poses a major public health threat, at least at the level of 2003 SARS^2^. A backtrend analysis suggested the. original basic reproduction number R0 to be about 2.4 to 2.5^3^ Li et al. Analyzed and predicted information such as demographic characteristics and patient exposure history. The estimated basic regeneration number R0 was 2.2 (95%CI,1.4 to 3.9), and the incubation period was 5.2 days (95% CI, 4.1 to 7). The number of confirmed cases doubles approximately every 7.4 days, and the interval from case onset to hospitalization and cure is 9.1 days (95% CI, 8.6-9.7)^4^. Shi Zhao et al. Estimated that the average basic regeneration number R0 ranges from 2.24 (95%CI: 1.96 to 2.55) to 3.58 (95% CI, 2.89 to 4.39)^5^. Wu et al. Analyzed and predicted by SEIR model, and estimated the basic regeneration number R0 to be 2.6 (95%CI, 2.47 to 2.86), and the estimated cumulative final confirmed cases were 75,815 (95% CI, 37304 to 130330), with approximately 6.4 confirmed cases Days (95% CI, 5.8 to 7.1) doubled^6^. Xiong et al. Predicted the basic regeneration number R_0_ at 2.985 through simulation of the EIR model. It is expected that the number of confirmed patients will reach its peak on February 16, 2020, and the number of confirmed patients will be 49,093^7^.

## METHODS

### MODIFIED SEIR MODEL

Compared with SARS, the symptoms of latent virus infection in latent patients are not obvious but they are as infectious as the confirmed patients. This feature brings great difficulties to the prevention and control of the epidemic. Because the classical SEIR model assumes that the infected person’s incubation period is not infectious, this assumption is quite different from the infection characteristics of the new coronavirus infection. Therefore, this article will use the revised SEIR model to analyze and predict the trend of the epidemic. The revised SEIR model still meets the classic assumptions: 1. The normal birth rate and mortality during the study period are not considered; 2. The influence of external factors on the model parameters is ignored; 3. The transmission method is person-to-person. The model divides the total population into four categories: susceptible populations (S, not yet infected but at risk of being infected), latent or exposed people (E, including people with mild or asymptomatic but infectious populations), Infected (I, confirmed to be infectious) and removed (R, including those who have been cured and died from neocoronavirus infection). The first case of New Coronavirus infection was found on December 8, 2019. The average incubation period of New Coronavirus infection was 7 days, so this article believes that the first patient infected with New Coronavirus may already exist on December 1, 2019. It has not been diagnosed yet, this article classifies those who have been infected with the new coronavirus but are not diagnosed as latent people; when the latent people are diagnosed as infected people, they will be treated in isolation in the hospital, then I think the infected people will no longer have the possibility of infecting others. Establish models using ordinary differential equations based on model assumptions(1)

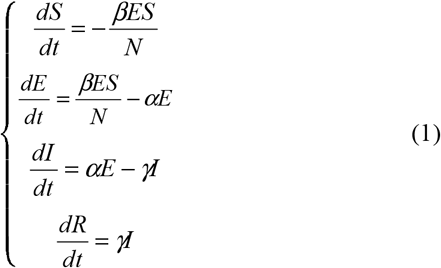

Where N represents the total population. N = S + E + I + R; βrepresents the probability of infection between the susceptible and the latent, and can also be expressed as the product of the average number of daily exposures of the latent (k) and the probability of being infected (b) That is, β= k * b, also known as the effective contact rate; αindicates the conversion rate of the latent to the infected, which is the reciprocal of the incubation period; γindicates the recovery rate, which is the reciprocal of the number of days in the treatment period.

Since the New Coronavirus infected person has a latent period and is also infectious during the incubation period, the state change rate in the SEIR model depends not only on the current state but also on the past state. The current number of infections in I(t) includes two parts. First, it was transformed by the latent person, but because of the incubation period of the new coronavirus infection, the study by Li et al. Estimated that the incubation period is 5.2 days (95% CI, 4.1 to 7). This article selects the incubation period as 7 days, so at time t The base for converting from a latent person to an infected person is the number of latent persons 7 days ago; the second is the number of infected persons at time t. Models can be built based on the delay difference equation(2)

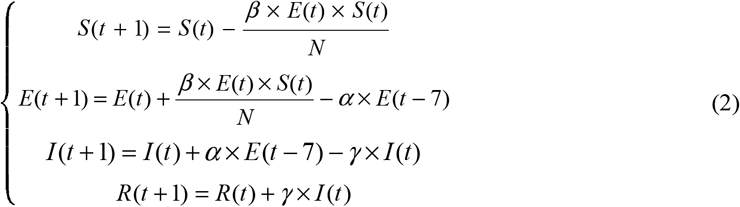

### ESTIMATION OF MODEL PARAMETERS

This article uses Wuhan as the main body of the simulation. The outbreak of the new crown virus infection is just before and after the Chinese Spring Festival. As of January 23, 2020, about 5 million people have flowed out of Wuhan. According to the 2018 statistical yearbook, the population of Wuhan is about 11.08 million. Therefore, this article assumes N = 6080000; if a susceptible person (S) comes in contact with a latent person (E) within a unit time, the susceptible person may be infected with the probability of infection and become a carrier (E). According to According to official reports, 1% to 5% of close contacts are diagnosed as infected, so the probability of contact between a susceptible population and a latent person is 1% to 5%. In this paper, 5% is selected as the infection probability b. The average daily number of close contacts of the latent person is k = 5, so the probability of infection βis 0.25; each exposed person (E) is transformed into the infected person (I) at a conversion rate α, and the incubation period refers to the time from infection to clinical symptoms of the disease. Covid-19 is currently estimated to have an incubation period of 1 to 12.5 days and a median of 5 to 6 days. According to information on other coronavirus diseases such as MERS and SARS, the incubation period for Covid-19 may be as long as 14 days. WHO recommends 14-day follow-up of contacts in confirmed cases. This article assumes that the incubation period is a maximum of 7 days. α= 0.143; the probability of each infected person (I) becoming a remover (R) is that during the early treatment process, because the treatment plan is designed to allow the infected person to survive 14 days, and achieve a healing effect through the autoimmune system; With the continuous improvement of the treatment plan, the treatment cycle has been significantly shortened. The treatment cycle selected in this paper is 9.1 days, so γ= 0.11. S (t), E (t), I (t), and R (t) are all greater than zero. The first patient was found on December 8, 2019, so the first case of latent appeared on December 1, 2019. December 1 is the 0th day, so S (0) = 6,079,999, E (0) = 1, I (0) = 0, and R (0) = 0.

## RESULTS

Using Matlab to simulate the model(2),you can get the figure 1.Day 0 in Figure 1 corresponds to the time when the first infection case (latency period) was found, that is, December 1, 2019. It can be seen from Figure 1 that the epidemic began to concentrate in about 79 days (ie, February 7, 2020). It can be seen that the number of infected people will reach its peak in about 89 days (February 28). In the case of quarantine precautions, the number of infected people may reach 2,384,803, and will eventually come to an end after 103 days (March 13

**Figure1.**
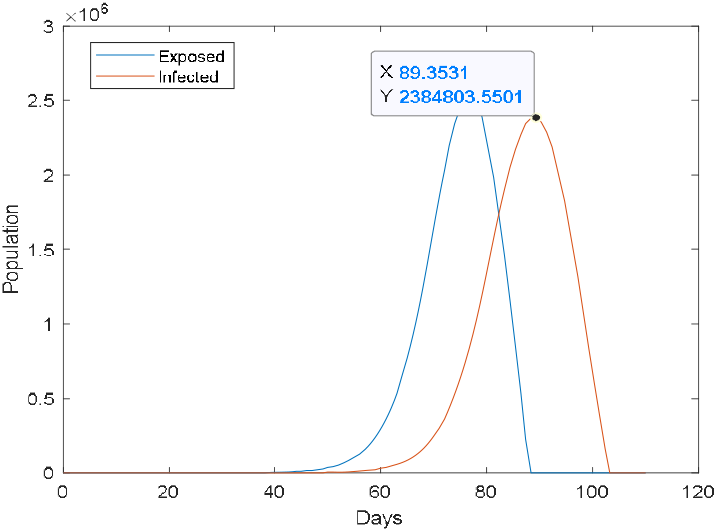
Modified SEIR model simulation results(k=5)

In fact, the Wuhan Municipal Government not only implemented the city closure measures on January 23, but also regulated traffic facilities in Wuhan and closed down most public places. We can simulate the intensity of government control by adjusting the average number of people k infected each day. From December 1st to January 23rd, the Wuhan Municipal Government did not take effective isolation measures, so when t <54, k = 5; when t≥ 54, the government adopted a series of strong measures such as the closure of the city and simulated them separately. The change in the number of infected persons at k = 1 and k = 2 can be obtained in Figures 2 and 3.

**Figure2.**
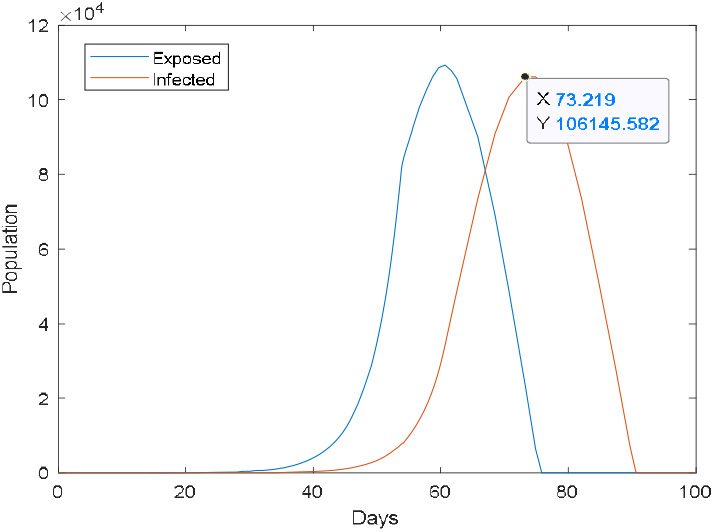
Modified SEIR model simulation results(t<54,k=5;t ≥54,k=2)

**Figure3.**
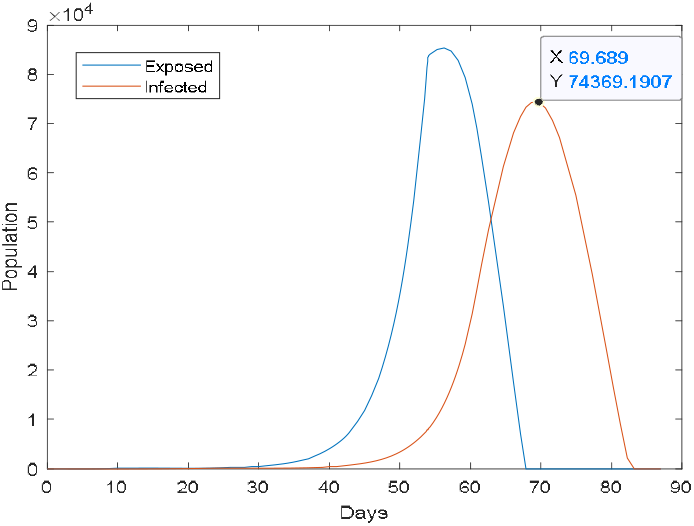
Modified SEIR model simulation results(t<54,k=5;t ≥54,k=1)

It can be seen from Figure 2 that when the government takes precautionary measures to control the average daily number of contacts per infected person k = 2, it can be seen that the number of infected persons reaches a peak after about 74 days, that is, February 12, 2020, and the number of infected persons is about 106,145 The epidemic situation is expected to come to an end in 90 days; when k = 1, it can be seen from Figure 3 that the number of infected people reached a peak about 70 days later, that is, February 8, 2020, and the number of infected people was about 74,369. It shows that when the government adopts effective quarantine measures, the number of infected people can be greatly reduced.

## DISCUSSION

The daily increase of newly diagnosed cases shows that the time point for Wuhan to adopt measures such as closing the city is somewhat delayed. Figures 4 and 5 respectively simulate the impact of closing the city one day in advance on the number of infected people. As can be seen from Figure 4, when Wuhan closed the city on January 22 a day in advance, the number of infected persons peaked after 73 days under the premise of k = 2, and the number of infected persons was about 86,252; under the premise of k = 1 It can be seen from Figure 5 that the number of infected persons peaked after about 69 days, and the number of infected persons was about 60,039. Compared with the closure on January 23, under the premise of k = 2, although the time to reach the peak of the number of infected persons has not changed much, the number of infected persons has decreased by 19,773; under the premise of k = 1, the infected persons The peak time was reached 2 days earlier, and the number of infected people decreased by approximately 14,330.

**Figure4.**
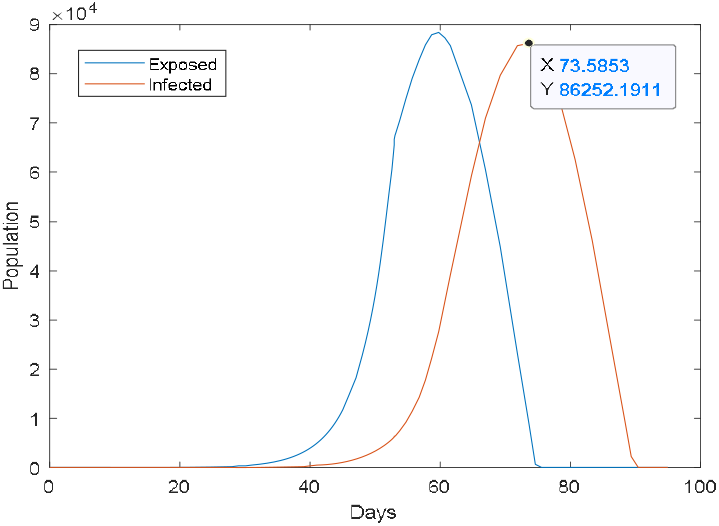
Modified SEIR model simulation results(t<53,k=5;t ≥53,k=2)

**Figure5.**
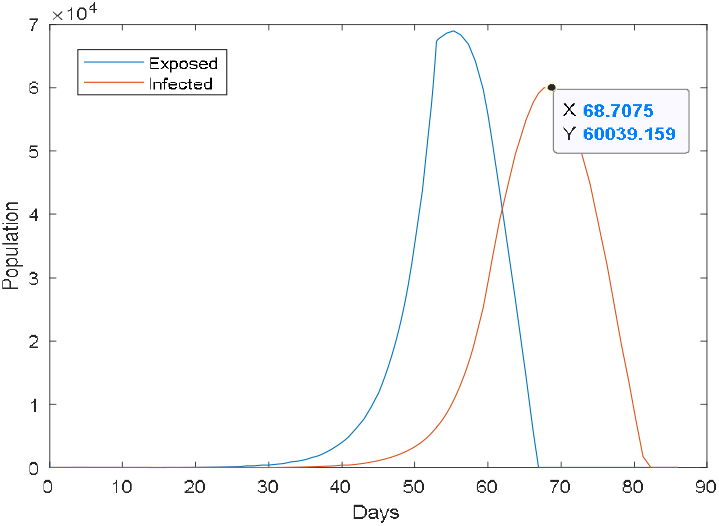
Modified SEIR model simulation results(t<53,k=5;t ≥53,k=1)

If the Wuhan municipal government lags behind for one day, it can be seen from the simulation results in Figures 6 and 7 that compared with the closure on January 23, the number of infected people will increase from 106,145 to 130,626 compared with the closure on January 23., An increase of 123%; under the premise of k = 1, the number of infected people will increase from 74,369 to 92,010, an increase of 124%

**Figure6.**
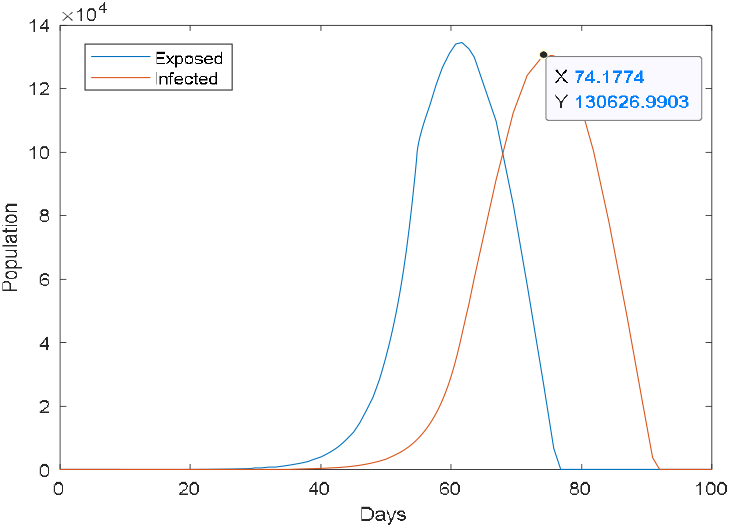
Modified SEIR model simulation results(t<55,k=5;t ≥55,k=2)

**Figure7.**
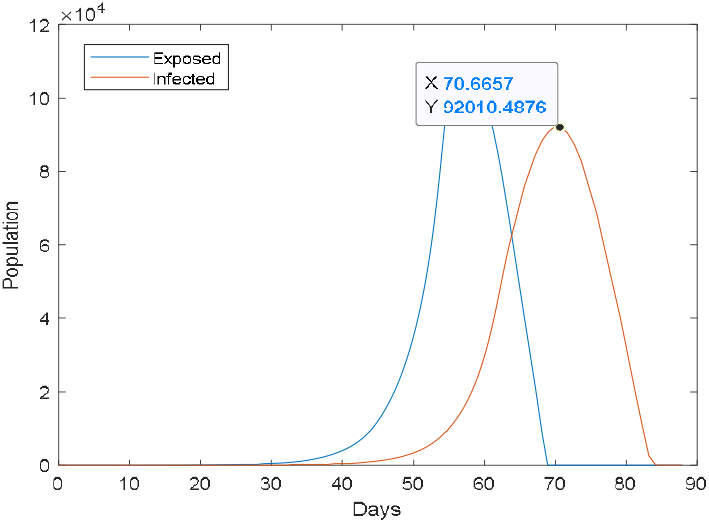
Modified SEIR model simulation results(t<55,k=5;t ≥55,k=1)

## CONCLUSIONS

Since January 23, 2020, all provinces across the country have adopted strict isolation and precautionary measures, and Wuhan has closed all public transportation facilities. China’s response to the epidemic is mainly divided into three stages: the first stage is to control the source of infection and block transmission around key areas in Hubei Province such as Wuhan; the second stage is to actively treat confirmed patients, reduce mortality, and prevent Export; the third stage is to reduce the epidemic situation, thoroughly control the disease epidemic, and significantly reduce the contact rate between people. In this paper, the trajectory of the curve is simulated by a time-lapse SEIR model. The model simulation shows that without any measures, the peak number of infected people will reach about 2,384,803 when the average daily number of contacts per infected person is k = 5; If Wuhan takes the measures of closing the city on January 22, 2020, the number of infected people will be reduced by about 19,773 on the premise of k = 2 compared with the closing of the city on January 23; The closure of the city on January 23 reduced 14,330 people. If Wuhan City is closed on January 24, the number of infected persons will increase by 123% under the premise of k = 2; the number of infected persons will increase by 124% under the premise of k = 1. It can be known that if the Wuhan Municipal Government takes strict precautionary measures in advance, the number of people infected will be greatly reduced. Therefore, local governments and public health departments should focus their efforts on controlling the source of infection and cutting off the transmission channels, such as early detection, early isolation, and early treatment, to reduce the gathering of large groups of people, and to regularly and completely kill public toilets and public spaces.

## Data Availability

All data generated or analyzed during this study are included in this article.

